# Parental Deprivation- and Threat-Based Factors Associated with Youth Emotion-Based Neurocircuitry and Externalizing Behavior: A Systematic Review

**DOI:** 10.1101/2022.08.10.22278633

**Authors:** Kathleen E. Feeney, Rosario Pintos Lobo, Megan M. Hare, Stephanie S.J. Morris, Angela R. Laird, Erica D. Musser

**Author notes:** **Corresponding Author:** Kathleen E. Feeney, Department of Psychology, Florida International University, Miami, FL, USA.

## Abstract

Parental factors, including negative parenting practices (e.g., family conflict, low monitoring), parental depression, and parental substance use, are associated with externalizing behaviors in youth; however, the mediating role of youth’s neurocircuitry in explaining these associations has been less studied. Both the dimensional and stress acceleration models provide frameworks for understanding how frontolimbic and frontoparietal networks implicated in emotional attention and regulation processes may be associated with parental factors. The current review builds upon this work by examining how deprivation- and threat-based parental factors are associated with youth externalizing behaviors through youth neurocircuitry involved in emotional functioning. A systematic review using PRISMA guidelines was completed and included five studies assessing parenting behaviors, six studies assessing parental depressive symptoms and/or diagnosis, and 12 studies assessing parental history of substance use. Overall, reviewed studies provided support for the dimensional and stress acceleration models within the context of deprivation and threat. There was limited support for the proposed mediation model, as only six studies tested for mediation. Specific recommendations for future work include more deliberate planning related to sample composition, improved clarity related to parental constructs, and consistency in methodology in order to better understand associations between contextual parental influences and youth neural and behavioral functioning.

---

Externalizing behaviors and disorders are associated with disruptions to academic, social, and emotional functioning (Bongers et al., 2008; Hinshaw, 1992; Kim et al., 2007). Extant literature has shown that parental psychopathology and parenting behaviors, as well as youth emotion regulation abilities, play a role in shaping the development of these behaviors (Batum & Yagmurlu, 2007; Pinquart, 2017; Schulz et al., 2021). Adverse parental experiences can be conceptualized broadly within the domains of deprivation (i.e., lack of nurturing environmental stimuli) and threat (i.e., presence of harmful stimuli; McLaughlin et al., 2014). Emerging empirical work (Hare et al., 2022) and a systematic review (McLaughlin et al., 2019) have begun to examine how deprivation- and threat-based parental factors are differentially associated with the structure, function, and connectivity of neural networks (e.g., frontolimbic, frontoparietal) underlying emotional processing and regulation. Expanding upon this work, the current study is the first to systematically review this literature and build a model describing associations among parental factors characterized by threat and/or deprivation, youth neural development, and externalizing behaviors.

## Externalizing Behaviors in Youth

Externalizing behaviors include aggression, defiance, hyperactivity, and impulsivity. These behaviors are most often observed among youth meeting Diagnostic and Statistical Manual of Mental Disorders, 5^th^ Edition (DSM 5) diagnostic criteria for attention-deficit/hyperactivity disorder (ADHD), conduct disorder (CD), and/or oppositional defiant disorder (ODD; Achenbach, 1966; Frick & Thorton, 2017). Externalizing behavior represents the most common cause of mental health referral among youth (Merikangas et al., 2011), with approximately 7.4% of youth in the United States receiving a diagnosis of an externalizing disorder (Ghandour et al., 2019). Across empirical and epidemiological studies, externalizing diagnoses are more common among male youth (King et al., 2018; Merikangas et al., 2009). Importantly, youth who exhibit externalizing behaviors are at elevated risk for delinquency, substance use, and violent behavior across development (Liu, 2004; Thompson et al., 2011). These outcomes are associated with high financial (e.g., legal fees, treatment) and societal (e.g., lower employment) costs (Foster et al., 2005; Knapp et al., 2011); thus, it is crucial to examine the pathways that lead to development of externalizing behaviors.

## Parental Factors and Youth Externalizing Behaviors

It is well established that parental factors, including parenting behaviors (e.g., family conflict, low monitoring, hostility), substance use, and psychopathology (e.g., depression) are associated with externalizing behaviors in youth (Beyers et al., 2003; Georgiou & Symeou, 2018; McKee et al., 2008; Shaw et al., 1994; Zhang et al., 2020). Both parental externalizing and internalizing (e.g., anxiety, depression) diagnoses have been associated with youth externalizing behaviors (McKee et al., 2008; McLaughlin et al., 2012). Further, Patterson’s coercion theory posits that coercive parent-child cycles (initiated by parental hostility and maintained by negative reinforcement) lead to antisocial and delinquent behaviors in youth (Patterson, 2016). One potential mediator of these associations is youth’s ability to regulate emotions (Morris et al., 2007). For example, parental hostility may lead to child difficulties with emotion regulation and consequently higher levels of externalizing behaviors (Siffert & Schwarz, 2011). Extant literature reviews suggest that parental factors may also affect youth neural structure, function, and connectivity involved in youth emotional functioning (Belsky & De Haan, 2011; Kerr et al., 2019); however, little work to-date has examined such associations among youth with externalizing behaviors, specifically.

## Conceptual Neurodevelopmental Pathway Models

Existing models of parental factors and youth emotion-linked neurodevelopment include the dimensional model and the stress acceleration model (McLaughlin & Sheridan, 2016; Callaghan & Tottenham, 2016). The dimensional model presents two separate dimensions of adverse parental influences: deprivation and threat. According to the dimensional model, deprivation is associated with alterations in the frontoparietal network (e.g., reduced cortical thickness and volume), which has been linked to executive functioning and attentional processes. In contrast, threat is associated with alterations in the frontolimbic network (e.g., reduced volume, elevated amygdala activation to threat) associated with emotion regulation.

A recent systematic review on deprivation (e.g., institutional rearing, neglect) revealed accelerated cortical thinning in the frontoparietal network (Colich et al., 2020). Additionally, adolescents with parental history of major depression (which may be characterized by disengagement or withdrawal; England et al., 2009) demonstrated lower dorsolateral PFC (dlPFC) activation to fearful faces compared to controls (Mannie et al., 2011), consistent with deprivation-related alterations in frontoparietal functioning. With respect to threat, early threatening parenting behaviors (e.g., maternal hostility, intrusiveness, negative affect) have been associated with reduced youth hippocampal subregion volumes (Blankenship et al., 2019), accelerated cortical thinning in the ventromedial prefrontal cortex (vmPFC; Colich et al., 2020), and increased amygdala activity while viewing angry and fearful facial stimuli (Pozzi et al., 2020), consistent with alterations in frontolimbic development (McLaughlin et al., 2019). Further, infants exposed to prenatal maternal depression (which, in addition to withdrawal, may be characterized by hostility; England et al., 2009) showed reduced connectivity between the amygdala and prefrontal regions (Posner et al., 2016). Interestingly, youth with family history of alcoholism (which may be characterized by abuse or neglect; Semidei et al., 2001) demonstrated lower amygdala volume and lower frontoparietal connectivity (Cservenka, 2016), consistent with both deprivation- and threat-related alterations. Thus, of note, while specific parenting behaviors may be easily characterized as deprivation or threat contexts, parents with depression and/or parents who engage in substance use may exhibit behaviors that can be categorized as both deprivation and threat (i.e., withdrawal, hostility; England et al., 2009; Semidei et al., 2001), accounting for the brain alterations consistent with predictions accounted for by the dimensional model.

In contrast to the dimensional model, the stress-acceleration model posits that exposure to adversity, primarily parental deprivation, may lead to rapid development of frontolimbic regions. For example, early maternal deprivation has shown associations with accelerated youth frontolimbic maturation (Gee et al., 2013; Silvers et al., 2016), consistent with this model (Callaghan & Tottenham, 2016). Similarly, infants exposed to prenatal maternal depression showed evidence of higher amygdala-vmPFC connectivity (Qiu et al., 2015). In terms of threat, exposure to maternal hostility during the preschool years has shown associations with negative amygdala connectivity with frontal areas during facial emotion tasks (e.g., sad expressions) in late childhood, consistent with patterns seen in adulthood (Kopala-Sibley, 2020). Similarly, youth with family history of alcoholism showed decreased amygdala activation (Cservenka, 2016). Thus, in some cases, both deprivation- and threat-related contexts have been associated with accelerated frontolimbic development, consistent with the stress-acceleration model.

A prior review on childhood adversity and neurodevelopment found evidence consistent with the dimensional model (i.e., frontoparietal alterations with deprivation, frontolimbic alterations with threat), but inconsistent evidence (i.e., support from only half of the studies) for the stress-acceleration model (McLaughlin et al., 2019). While there was stronger evidence for accelerated development of neural structure, the findings on amygdala-mPFC connectivity were mixed, possibly due to variability in sample sizes and the nature of the specific tasks completed. Notably, a more recent meta-analysis and systematic review did not find consistent associations between early life adversity involving threat or deprivation and amygdala-PFC connectivity (Colich et al., 2020). Of importance, contextual parenting experiences are not deterministic (Thompson, 2016), prompting the need for more work to identify which youth are more prone to frontolimbic and frontoparietal disruption, so that interventions and prevention efforts may be more targeted. Thus, the current review will aim to identify patterns in brain structure and function that occur across studies and highlight any divergences which may be related to diversity of samples or tasks.

## Youth Emotional Neurocircuitry and Externalizing Behaviors

Both structural and functional brain differences within emotion-based circuits, including those implicated in both the dimensional model and stress acceleration model, have been observed among youth exhibiting externalizing behaviors. For example, externalizing behaviors in typically developing youth, even at subclinical levels, have been associated with reduced amygdala and prefrontal cortex (PFC) volume (Caldwell et al., 2015; Montigny et al., 2013) and reduced hippocampal volume (Bos et al., 2018). In terms of functional activation, externalizing behavior in youth has been associated with hypoactivity in the medial PFC (mPFC) prior to risky decision-making and in the insula during facial emotion processing (e.g., happy expressions) among female youth (Crowley et al., 2017; Whittle et al., 2015).

With respect to functional connectivity, youth have shown lower white matter integrity in a connective tract between limbic and frontal regions (Andre et al., 2020a) and specifically, male youth with externalizing diagnoses have shown decreased connectivity between frontal and limbic regions (Shannon et al., 2009). Further, youth in middle childhood exhibiting higher levels of aggressive behavior have shown higher global functional connectivity within the amygdala and reduced frontolimbic connectivity at rest in comparison to controls (Sukhodolsky et al., 2021). Additionally, Ameis and colleagues (2014) found stronger coupling between the amygdala and vmPFC in adolescents exhibiting lower, but not higher, externalizing behaviors. In sum, the literature supports the notion that youth with externalizing behaviors exhibit atypical structural (i.e., reduced volumes and white matter) and functional (i.e., weaker frontolimbic connectivity) development in areas linked to emotional functioning and areas implicated in both the dimensional model and stress acceleration model. While these patterns appear to be better accounted for by the dimensional model, it will be important to examine the pathways to these youth externalizing behaviors from parental factors characterized by deprivation and threat, as this may account for heterogeneity in neurocircuitry among youth.

## Current Review

Prior work suggests divergent neurodevelopmental pathways exist based upon adverse parental influences characterized by deprivation and/or threat; however, there have been varied findings due to inconsistencies in samples and tasks. Additionally, studies to date have not yet comprehensively examined how these neurodevelopmental pathways impact youth externalizing behavioral outcomes, despite work that has shown alterations in neurodevelopment (i.e., within the frontolimbic network) that may lead to youth psychopathology (Jones et al., 2017). The current review will build upon prior work by examining how parental factors characterized by deprivation and/or threat contexts are associated with youth neural structure and function (e.g. activation and connectivity) and consequently youth externalizing behaviors. We will present a conceptual model whereby youth neurocircuitry serves as a mediator between parental factors and externalizing behaviors (see Figure 1), which builds on the dimensional model and stress acceleration model (McLaughlin & Sheridan, 2016; Callaghan & Tottenham, 2016). Thus, this review will serve to improve the understanding of heterogeneity in youth neurocircuitry, as well as the etiology of externalizing behaviors, through the application and extension of preexisting models based on deprivation vs. threat contexts. An earlier review by Belsky and de Haan (2011) on parenting and youth neurodevelopment noted that this area of study was “not even yet in its infancy”; thus, the current review aims to synthesize the still limited, but emergent, work in this area. Based on prior studies, we expect to find evidence supporting both the dimensional model and the stress acceleration model. Further, as shown in the proposed model, we hypothesize that deprivation- and threat-related parenting factors will be associated with elevated externalizing behavior through altered neural structure and function (whether indicative of accelerated or delayed development).

**Figure 1.**
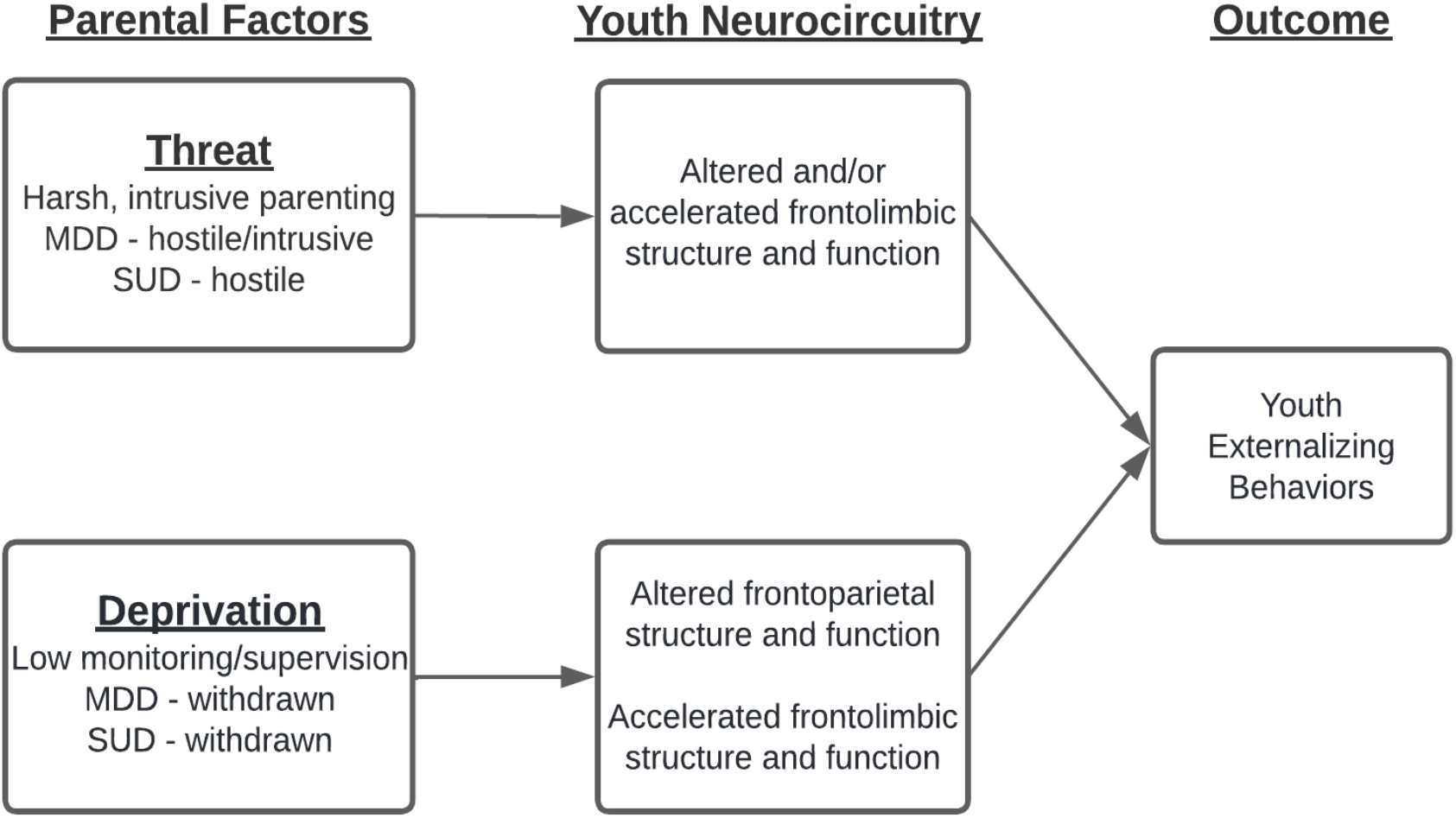
Conceptual Mediation Model of Neurodevelopment.

## Methods

### Search Procedures

For the current review, PRISMA reporting guidelines for a systematic review were used (Moher et al., 2009; see Figure 2). Two researchers (initials masked for review) completed two independent database searches and titles and abstracts were screened to determine eligibility (92.3% agreement). A third reviewer (initials masked for review) reconciled disagreements (two articles were confirmed for inclusion by consensus in this manner). The following search terms were utilized in PubMed and PsycInfo: (1) “((externalizing) AND parent*) AND ((fMRI) OR (functional MRI) OR (functional magnetic resonance imaging))”, (2) “((externalizing) AND parent*) AND ((EEG) OR (electroencephalogra*))”, (3) “((externalizing) AND parent*) AND emotion* AND neuro*”, and (4) “((externalizing) AND parent*) AND emotion* AND neural”.

**Figure 2.**
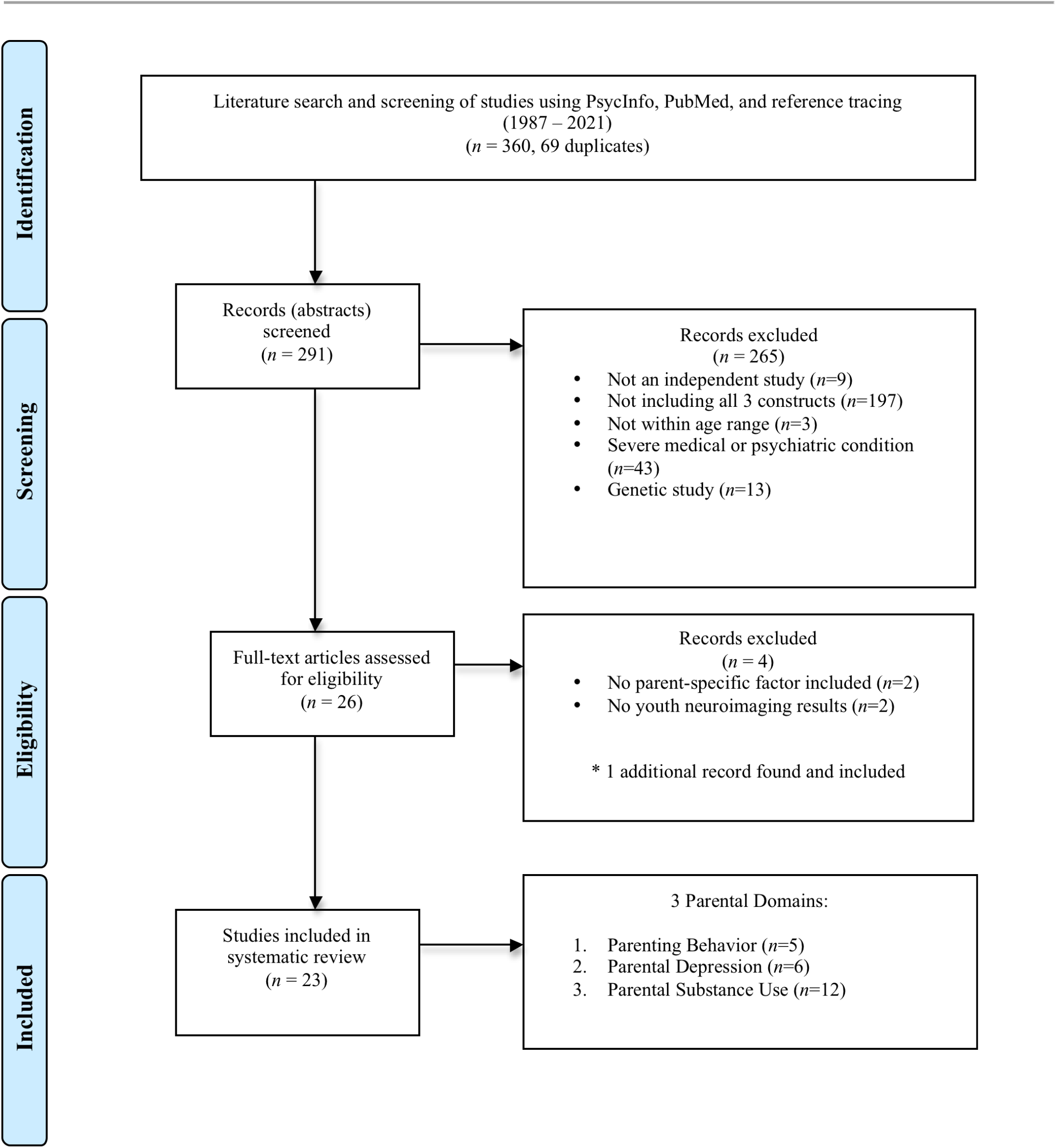
<u>PRISMA Flowchart for identification and eligibility of articles.</u> Template provided by (Moher et al., 2009).

### Selection Criteria

A large developmental period (preschool age [3 years] through young adulthood [25 years]) was assessed in this review. Articles needed to contain all three constructs of inquiry: at least one parental factor (i.e., parenting behavior, parental depression, or parent substance use), differences in functional and/or structural youth emotion-based neurocircuitry, and youth externalizing behaviors for inclusion. Studies that focused exclusively on specific genetic (i.e., biomarkers) and hormonal influences, as well as studies that did not include a youth neuroimaging component, were excluded. Additionally, articles were excluded if the participant population included youth with autism, brain tumors, epilepsy, head injury, developmental delay, intellectual disability, and/or tic disorders, as these conditions may present confounds when considering neural development.

### Data Extraction

The following data was extracted from the chosen articles: (1) authors and year of publication, (2) sample characteristics (youth age, biological sex, ethnicity, race), (3) sample composition (e.g., clinical or community sample), (4) method of neuroimaging data acquisition, (5) task completed during EEG or functional MRI and/or whether only structural MRI was acquired, (6) neuroimaging results related to youth brain structure and function, (7) parent-report and observational measures of parental factors, and (8) youth behavioral outcomes.

## Results

Out of the 291 total articles identified, which were published between 1987 and 2021 (excluding overlap across databases; 360 total articles with 69 duplicates), 26 articles were selected following the initial screening of titles and abstracts. Of the 265 articles that were deemed not eligible, 197 did not include all three constructs (parenting, youth neurocircuitry, and youth externalizing behavior), 43 met criteria for a medical/psychiatric exclusion (e.g., autism), 13 were genetic studies, nine were not independent empirical studies (e.g., meta-analysis, review), and three studies involved participants outside the target age range. After a full article review for inclusion, two articles were excluded due to the lack of a parent-specific factor (e.g., family history rather than parental history) and two articles were excluded due to lack of youth neuroimaging results. One additional article was found via reference tracing and was included due to meeting inclusion criteria for this review. Overall, 23 articles met inclusion criteria and were organized by three domains: parenting behavior (n=5), parental depression (n=6), and parental substance use (n=12). See Figure 2 for a detailed outline of inclusion for the reviewed studies in accordance with PRISMA guidelines and Table 1 for detailed study information (e.g., brief demographics, methods, results).

**Table 1.** Overview of Reviewed Articles.

| Study | Participants |  |  | Task | Results |  |
| --- | --- | --- | --- | --- | --- | --- |
|  | *N* | Age | Sample Details |  | Behavioral Associations | Brain-Behavior Associations |
| Parenting Behavior |  |  |  |  |  |  |
| Barbosa et al., 2018 | *N* = 88 | *M* = 9.42, *SD* = 1.08 years | Healthy youth; early and late pubertal development | Task-based fMRI: facial expressions task (angry, calm, fearful, happy) | ↑ inconsistent discipline, ↑ child externalizing behavior. | ↑ poor monitoring and supervision, ↓ vIPFC activation to fearful > calm.<br>↑ corporal punishment, ↓ vIPFC activation (late females); ↑ corporal punishment, ↑ vIPFC activation (late males) to fear>calm.<br>↑ corporal punishment, ↓ dIPFC, frontal pole, anterior insula activation (late females) and ↓ frontal pole (early males); ↑ corporal punishment, ↑ dIPFC, frontal pole, anterior insula activation (late males).<br>No significant mediation. |
| Chaplin et al., 2019 | *N* = 66 | 12-14 years; *M* = 12.59, *SD* = .70 years | Community sample | Task-based fMRI: IAPS-Negative, Neutral, Positive |  | **Females:** ↑ maternal negative parenting, ↑ activation in R ACC to negative emotional stimuli.<br>↑ activation in L anterior insula to negative stimuli, ↑ adolescent substance use.<br>**Males:** ↑ maternal negative parenting, ↓ activation in L and R anterior insula and L ACC to negative stimuli.<br>No associations between brain activity and externalizing symptoms. |
| Demers et al., 2019 | *N* = 88 | *M* = 30 years; recruited at age 6-12 years | Child maltreatment history (*N*=48) | sMRI | ↑ maternal relationship quality, ↓ externalizing symptoms. | ↑ maternal relationship quality, ↑ frontal lobe volume (non-maltreated).<br>No associations between brain function and adult externalizing symptoms. |
| Holz et al., 2018 | *N* = 172 | Assessed from age 6 | High and low familial risk for | Task-based fMRI: Reward anticipation and delivery; | ↑ family risk of psychopathology, ↓ | ↑ familial risk, ↓ caudate head activity (anticipation). |
|  |  | months to 25 years | psychopathology | contrast of anticipation - monetary vs. verbal and contrast of delivery - win vs. no win | maternal stimulation, ↑ psychopathology (ADHD, ODD, etc.).<br>↑ maternal stimulation, ↓ ADHD (↑ familial risk). | ↑ familial risk, ↑ caudate head activity (delivery).<br>↑ maternal stimulation, ↑ caudate head activity (high familial risk);<br>↑ maternal stimulation, ↓ caudate head activation (no/low risk).<br>↑ maternal stimulation → ↓ caudate head activity (delivery) → ↓ ADHD. |
| Shackman & Pollak, 2014 | *N* = 50 | 7-9 years | All male sample; Maltreatment history (*N*=17), Control (*N*=33) | EEG: ERP during emotion recognition task (emotional oddball and provocation task) | ↑ physical maltreatment (vs. no maltreatment), ↑ child negative affect, ↑ child aggressive behavior. | ↑ physical maltreatment → ↑ P3b amplitude (i.e., ↑ attention) to angry faces → ↑ child negative affect.<br>P3b amplitude unrelated to externalizing behaviors. |
| **Parental Depression** |  |  |  |  |  |  |
| Ashman et al., 2008 | *N* = 159 dyads | Assessed from infancy to 6.5 years | Children of mothers with and without depression | EEG: EEG power while watching video clips – baseline, happy, sad, neutral, anxious/scared | ↑ maternal depression, ↑ child externalizing behavior (aggression and ADHD).<br>↓ but stable and decreasing maternal depression (vs. no depression), ↑ child externalizing ADHD. | ↑ maternal depression, ↑ EEG alpha power (↓ frontal brain activation).<br>No association between frontal brain activation and externalizing behaviors. |
| Dawson et al., 2003 | *N* = 124 mothers and their child | Child: *M*<sub>age</sub> = 3.5 years | History of maternal depression (*N*=65) | EEG: at rest and during emotion-eliciting stimuli presentation | ↑ maternal depression, ↑ marital discord and stress, ↑ child behavior problems. | ↑ maternal depression, ↓ frontal brain activation.<br>↓ frontal and parietal brain activation, ↑ behavior problems.<br>↑ maternal depression → ↓ frontal brain activation → ↑ behavior problems. |
| Forbes et al., 2006a | *N* = 57 | 3-9 years | Parents with child onset depression or COD (*N*=41) | EEG: disappointment task | No differences in externalizing behaviors between COD and control subjects. | ↑ relative left frontal activity, ↑ externalizing problems (COD).<br>No mediation conducted. |
| Forbes et al., 2006b | *N* = 74 mother-child dyads | 3-9 years | History of maternal COD (*N*=44) | EEG: resting state | ↑ maternal depression, ↑ child aggressive behaviors. | ↑ left frontal asymmetry, ↑ aggressive child behavior (COD).<br>Males: Left frontal asymmetry (vs. right frontal asymmetry), ↑ child aggressive behaviors. |
| Qu et al, 2016 | *N* = 23 | *M* = 15.78, *SD* = .6 |  | Task-based fMRI: BART risk-taking/reward task | ↑ parental depressive symptoms, ↑ adolescent | ↑ parental depressive symptoms, ↑ ventral striatum and dlPFC |
|  |  | years |  |  | risk-taking and externalizing behaviors. | activation over time to reward during risk-taking.<br>↑ parental depressive symptoms<br>→ ↑ longitudinal VS activation<br>→ ↑ adolescent externalizing behavior. |
| Wang et al., 2020 | *N* = 183 | 4.5 years |  | sMRI; rs-fMRI |  | **Females:** ↑ maternal depressive symptoms, connectivity between L/R vmPFC & L medial prefrontal thalamus, right subcallosal cortex & L vmPFC/R ACC; Connectivity between R lat. amygdala & L ACC, R vmPFC & R med. amygdala, ↑ externalizing behaviors.<br>**Males:** ↑ maternal depressive symptoms, connectivity between R vmPFC & L/R ACC, L OFC & L ventral caudate; ↑ externalizing behaviors, connectivity between L med. prefrontal thalamus & bilateral ACC, L occ. thalamus & left lat. OFC, L vm putamen & L d. caudate. |
| **Parental Substance Use** |  |  |  |  |  |  |
| Andre et al., 2020b | *N* = 66 | 7-16 years | PAE + adverse postnatal exposure to maltreatment, PAE only, age- and gender-matched controls | sMRI; DTI | ↑ prenatal alcohol exposure (regardless of maltreatment), ↑ child externalizing behavior. | ↑ prenatal alcohol exposure (no postnatal adversity), ↓ FA in bilateral cingulum and left uncinate fasciculus, ↓ L ACC volume. No brain associations with externalizing behaviors. |
| Carlson & Iacono, 2008 | *N* = 369 | Assessed from age 17 to 24 years | Severe (*N*=82): paternal alcohol use + antisocial behavior; Intermediate (*N*=126): alcohol dependence only; Low | EEG: ERP during rotated heads task |  | ↑ paternal alcohol use + antisocial behavior, ↓ initial P300 value and less P300 change over time. ↓ initial P300 value, ↑ child externalizing and early-onset substance use disorders. |
|  |  |  | (*N*=161): no substance use or disorder |  |  |  |
| Ehlers et al., 2001 | *N* = 104 | 7-13 years | At least one biological parent met for lifetime history of AUD | EEG: ERP during facial discrimination task |  | ↑ parental substance use, ↓ frontal P3 amplitude.<br>↓ P3 amplitudes in posterior areas, ↑ delinquent behaviors. |
| Euser et al., 2013 | *N* = 193 | 12-20 years | High risk (*N* = 83): parental history of SUD; Low risk (*N*=110): control | EEG: visual novelty Oddball paradigm | ↑ parental history of SUD, ↓ parental emotional warmth. | ↑ parental substance use, ↓ novelty-P300 amplitude above and beyond all else.<br>↓ target-P300 amplitude, ↑ externalizing behavior. |
| Heitzeg et al., 2008 | *N* = 28 | 16-20 years | COA + high problem drinking (*N*=11); COA + low problem drinking (*N*=11); No parental history AUD or problem drinking (*N*=6) | Task-based fMRI: positive, negative, neutral valence word viewing task | Parental history of AUD + ↑ drinking problems, ↑ child externalizing behavior. | COA + ↓ problem drinking: ↑ bilateral OFG and ↑ left insula/putamen to emotional stimuli.<br>COA + ↑ drinking problem: ↑ dmPFC, ↓ ventral striatum and extended amygdala bilaterally to emotional stimuli. |
| Heitzeg et al., 2010 | *N* = 61 | 16-22 years | Parents with AUD - low and high alcohol problems (*N*=41); No AUD diagnosis (*N*=20) | Task-based fMRI: Go/no-go task | ↑ parental history of AUD (regardless of current drinking problems), ↑ externalizing behaviors. | ↓ alcohol use (regardless of family AUD history), ↓ orbital and left medial prefrontal activation, ↑ alcohol use, ↑ mPFC activation continued.<br>↑ (Failure to deactivate) ventral caudate activity to successful inhibition trials, ↑ externalizing behaviors. |
| Heitzeg et al., 2014 | *N* = 45 | 9-12 years | High parental AUD (*N*=26 with 13 problem users and 13 nonusers); Controls (*N*=19) | Task-based fMRI: Event-related go/no-go task | ↑ problem use + parental AUD history, ↑ child externalizing behaviors. | ↑ parental AUD history, ↓ left MFG activation, ↑ externalizing behavior.<br>Parental AUD + problem use: ↓ left MFG activation during failed inhibition compared to non-users.<br>↑ left MFG activation predicted group membership over and above externalizing problems. |
| Holla et al., 2019 | *N* = 140 | 8-23 years | Parental AUD history + substance-naïve (*N*=75); Age- | Resting-state sMRI | ↑ parental AUD history, ↑ child externalizing behavior. | Adolescence: ↑ parental AUD history, ↑ cortical thickness in bilateral precentral gyri, left caudal middle frontal gyrus, bilateral |
| | | | matched substance-naïve male controls ($N=65$) | | | temporo-parietal junction, left inferior-frontal gyrus, right inferior-temporal gyrus. Group differences gone by young adulthood.<br>↑ cortical thickness in left MFG and inferior-parietal lobule, ↑ externalizing symptoms. |
| Hulvershorn et al. 2013 | $N = 37$ | 10-14 years | Non-substance using fathers with past or current SUD, ADHD+DBD ($N=19$); Healthy controls ($N=18$) | Task-based fMRI: block-design task; facial expression matching | ↑ paternal SUD history, ↑ emotional lability/negativity, ↓ emotion regulation. | ↑ paternal SUD history, ↑ mPFC activation. |
| Kamarajan et al., 2015 | $N=1864$ | 12-25 years | Family history of alcoholism ($N=1569$); Community sample ($N=295$) | EEG: ERP during monetary gambling task (gain and loss conditions) | ↑ family history of alcoholism, ↑ impulsivity, ↑ externalizing disorders. | ↑ family history of alcoholism [older (16–25 years) male and younger (12–15 years) female subgroups], ↓ P3 amplitude. ↑ family history of alcoholism (males) during the loss condition, ↓ P3 amplitude. ↑ P3 amplitude at anterior sites and ↓ at posterior areas were seen in older compared to younger participants. ↑ family history of alcoholism, ↓ CSD activity during both loss and gain conditions at frontal regions. |
| Kwon et al., 2021 | $N = 90$ | 11-12 years | All participants had ADHD and DBDs | fMRI: resting-state | | ↑ family history of substance use, ↑ functional connectivity between the right lateral OFC seed and anterior cingulate cortex. ↑ family history of substance use, ↑ negative functional connectivity between the left lateral OFC seed and right postcentral gyrus and between the left NAcc seed and right middle occipital gyrus. ↑ negative connectivity between right occipital/left NAcc (↑ family substance use history), ↓ emotion regulation. |
| Kwon et al., 2021 | N = 90 | 11-12 years | All participants had ADHD and DBDs | fMRI: resting-state |  | ↑ family history of substance use,<br>↑ functional connectivity between the right lateral OFC seed and anterior cingulate cortex.<br>↑ family history of substance use,<br>↑ negative functional connectivity between the left lateral OFC seed and right postcentral gyrus and between the left NAcc seed and right middle occipital gyrus.<br>↑ negative connectivity between right occipital/left NAcc (↑ family substance use history), ↓ emotion |

*Note.* ACC = anterior cingulate cortex, ADHD = attention-deficit/hyperactivity disorder, AUD = alcohol use disorder, COA = child of alcoholic, COD = child onset depression, CSD = cortical spreading depression, DBD = disruptive behavior disorder, dlPFC = dorsolateral prefrontal cortex, dmPFC = dorsomedial prefrontal cortex, EEG = electroencephalogram, ERP = event-related potential, FA = fractional anisotropy, fMRI = functional magnetic resonance imaging, IAPS = International Affective Picture System, MFG = middle frontal gyrus, mPFC = medial prefrontal cortex, NAcc = nucleus accumbens, OFC = orbitofrontal cortex, PAE = prenatal alcohol exposure, sMRI = structural magnetic resonance imaging, SUD = substance use disorder, vlPFC = ventrolateral prefrontal cortex, VS = ventral striatum, ↑ = higher/increased, ↓ = lower/decreased, → = testing of a mediation pathway.

### Parenting Behavior, Emotion Circuitry, and Youth Externalizing Behavior

Four of the five studies examining parenting behavior included in this review reported behavioral findings consistent with extant literature (regardless of being deprivation- or threat-related), such that inconsistent discipline, low maternal stimulation, and physical maltreatment were associated with elevated externalizing behaviors, while higher maternal relationship quality and high maternal stimulation were associated with reduced externalizing behaviors (Barbosa et al., 2018; Demers et al., 2019; Holz et al., 2018; Shackman & Pollak, 2014).

When examining structural brain differences associated with threat-type parental factors (e.g., child maltreatment including abuse, criticism, and corporal punishment), one study found that higher maternal relationship quality in adolescence was associated with higher frontal lobe volume in adulthood, but only among youth who did not have a child maltreatment history (Demers et al., 2019). Of note, child maltreatment in this study included emotional, physical, and sexual abuse, as well as neglect, and was not necessarily inflicted by a parent. With respect to functional differences, physical maltreatment by a parent was associated with increased P3b amplitude (i.e., attention) to angry faces; however, P3b amplitude was not associated with externalizing behaviors in this study, and thus, mediation was not supported (Shackman & Pollak, 2014). Further, the effects of corporal punishment among youth (Barbosa et al., 2018) and maternal negative parenting behaviors among adolescents (e.g., criticism, harsh tone; Chaplin et al., 2019) on functional activation varied by biological sex and pubertal timing. The effects of corporal punishment and negative parenting behaviors generally resulted in lower frontal activation for females (i.e., vlPFC) and early developing males (i.e., frontal pole), higher limbic activation for females (i.e., right ACC, left anterior insula), as well as higher frontal activation (i.e., dlPFC, frontal pole) and lower limbic activation (i.e., left ACC, left and right anterior insula) for late developing males. However, in these studies, significant mediation pathways from parenting behaviors to youth externalizing behavior through brain functioning were not observed.

In contrast, poor parenting practices more closely related to deprivation (e.g., low monitoring and supervision) were associated with lower vlPFC activation to negative (e.g., fearful) compared to neutral (e.g., calm) facial stimuli among youth (Barbosa et al., 2018). However, again, significant mediation pathways from parenting behaviors to youth behavior through brain functioning were not supported in this study. In contrast, when assessing for maternal stimulation, which could be considered a positive counterpart to neglect, lower caudate activity mediated the association between high maternal stimulation and lower symptoms of ADHD (Holz et al., 2018). It is important to note, however, that this was just one study and thus results should be interpreted cautiously. Taken together, there was support for the dimensional model for both deprivation and threat contexts, as well as the stress acceleration model with respect to the threat context only; however, the proposed mediation model was not supported. Interestingly, there was also evidence of disruption in frontoparietal networks in the threat context.

### Parental Internalizing Influences on Youth Neural Circuitry and Behavior

Parental depressive symptoms reported across four out of six studies were associated with youth externalizing behaviors (Ashman et al., 2008; Dawson et al., 2003; Forbes et al., 2006b; Qu et al., 2016). Two reviewed studies appeared to support the proposed mediational threat pathway (see Figure 1) as youth frontolimbic activation was a mediating factor between parental psychopathology and youth externalizing behavior. Specifically, associations between parental depressive symptoms and elevated externalizing behavior were mediated both by higher ventral striatum activation (Qu et al., 2016) and lower frontal activation (Dawson et al., 2003). Additional work examining children of parents with depression showed that these youth exhibited low frontal activation during emotion-eliciting stimuli presentation (Ashman et al., 2008), as well as left frontal asymmetry at-rest (Forbes et al., 2006b) and during a disappointment task (Forbes et al., 2006a) compared to peers. In contrast to the study by Dawson and colleagues (2003), low frontal activation was not associated with externalizing behavior in a later study (Ashman et al., 2008). Left frontal asymmetry was associated with externalizing behavior; however, mediation was not assessed (Forbes et al., 2006a; Forbes et al., 2006b).

Similarly to threat-based parenting factors, sex differences were observed in the context of maternal depressive symptoms such that the dorsal portion of the mPFC (i.e., dorsal ACC) in males and ventral portion of the mPFC (subcollosal cortex and vmPFC) in females were associated with both maternal depressive symptoms and youth behavioral problems (Wang et al., 2020). More specifically, females exhibited higher amygdala-vmPFC connectivity, whereas males exhibited lower activation in reward networks. With respect to parental depression, there was support for the dimensional model in the threat context, as well as the stress acceleration model in either the context of deprivation or threat. Further, the proposed mediation model was supported in the threat context.

### Parental Externalizing Influences on Youth Neural Circuitry and Behavior

Prenatal alcohol exposure and parental history of alcohol use disorder were both associated with youth externalizing behaviors across studies (Andre et al., 2020b; Heitzeg et al., 2008; Heitzeg et al., 2010; Heitzeg et al., 2014; Holla et al., 2019; Kamarajan et al., 2015; Venkatasubramanian et al., 2007). Parental history of substance use disorder was associated with higher child negative emotionality and lability, as well as lower emotion regulation (Hulvershorn et al., 2013). Mediation was not assessed in any studies assessing for parental substance use.

Youth who had parents with alcohol use disorders demonstrated higher cortical thickness in frontal regions (i.e., delay in cortical pruning; Holla et al., 2019) and reduced P300 amplitudes (i.e., lower cognitive engagement; Carlson & Iacono, 2008; Ehlers et al., 2001; Euser et al., 2013; Kamarajan et al., 2015). Similarly, some work found that youth from middle childhood to adolescence (Andre et al., 2020b) and adulthood (Venkatasubramanian et al., 2007) with parental substance use history had smaller frontal and limbic volumes (e.g., ACC, corpus callosum, cingulum, genu, and isthmus) and exhibited lower frontal activation (Heitzeg et al., 2014).

In contrast, other studies were fairly consistent in showing that high-risk youth (i.e., parental history of substance use disorder) in middle childhood (Hulvershorn et al., 2013; Kwon et al., 2021) through adolescence (Heitzeg et al., 2008; Heitzeg et al., 2010; Hulvershorn et al., 2013) exhibit heightened activation in prefrontal regions and reduced activation in limbic regions (i.e., striatum, amygdala) compared to controls across tasks. Thus, with respect to parental substance use, there was support for the dimensional model in the context of deprivation and threat, as well as the stress acceleration model in the context of either deprivation or threat. While mediation was not tested, there were significant associations between lower frontal activation and heightened limbic activation with externalizing behaviors. Thus, there may be potential support for the proposed mediation model in a deprivation or threat context.

## Discussion

Across healthy and high-risk (e.g., history of maltreatment) samples, parental factors were associated with youth behavioral outcomes in the expected directions (See Table 1 for study results). The model presented within this review (see Figure 1) expanded upon pre-existing models by distinguishing two pathways by which threat- and deprivation-related factors may result in elevated youth externalizing behaviors through altered functioning in frontolimbic and frontoparietal networks. As described previously, the dimensional model proposed deficient neural development, while the stress acceleration model proposed accelerated neural development due to parental factors.

Across nine studies, there was support for the deprivation-based dimensional model with disruption in frontoparietal regions (i.e., lower activation, reduced cortical pruning) due to poor monitoring and supervision, maternal depression, as well as parental substance use (Ashman et al., 2008; Barbosa et al., 2018; Carlson & Iacono, 2008; Dawson et al., 2003; Ehlers et al., 2001; Euser et al., 2013; Heitzeg et al., 2014; Holla et al., 2019; Kamarajan et al., 2015). When examining maternal stimulation (i.e., low deprivation; Holz et al., 2018), a mediation model was supported such that higher maternal stimulation led to lower ADHD symptoms through reduced caudate head activity. This suggests evidence for protective factors within the mediated pathway, although it is limited to one study. With respect to parental depression, Dawson and colleagues (2003) conducted the only study to report parental behaviors related to depression within the sample and noted higher levels of withdrawn behavior consistent with deprivation. While parents with depression may engage in hostile or withdrawn parenting or both (NRC and IM, 2009), parental neglect may be a more salient mediator than abuse in the association between parental depression and youth outcomes (Mustillo et al., 2011). The mediation model presented in this review was supported during toddlerhood by one study assessing for parental depression (Dawson et al., 2003), but was not supported during school age by a study assessing for poor monitoring and supervision (Barbosa et al., 2018) or a different study assessing for parental depression (Ashman et al., 2008). Studies assessing for parental substance use did not specifically test for mediation but noted significant associations between lower brain activation or reduced cortical pruning and externalizing behavior (Carlson & Iacono, 2008; Ehlers et al., 2001; Euser et al., 2013; Heitzeg et al., 2014; Holla et al., 2019); thus, these implied mediation pathways should be more closely examined in future studies.

The threat-based dimensional model was supported across nine reviewed studies, such that corporal punishment, negative parenting behaviors, maternal depressive symptoms, and parental history of substance use led to disruptions (i.e., smaller volumes, lower frontal and higher limbic activation) in frontolimbic regions (Andre et al., 2020b; Barbosa et al., 2018; Chaplin et al., 2019; Heitzeg et al., 2008; Heitzeg et al., 2010; Heitzeg et al., 2014; Qu et al., 2016; Venkatasubramanian et al., 2007; Wang et al., 2020). The proposed mediation model was not supported in studies assessing for corporal punishment, negative parenting, and parental substance use. However, it was supported in a study examining the association between parental depressive and adolescent externalizing symptoms through prolonged ventral striatum activation (Qu et al., 2016), consistent with the mediational threat pathway. Notably, this study recruited a community sample, suggesting that even mild to moderate levels of depressive symptoms may contribute to abnormal limbic activation and externalizing behavior. While this mediation model fell more in line with the threat-based pathway, data collection of depressive symptoms and diagnoses did not allow for parsing whether specific symptoms observed in the sample were more threat- or deprivation-based. Further, disrupted frontolimbic structure and function was associated with externalizing behaviors across studies examining a threat context, consistent with the proposed mediational threat pathway; however, full mediation was not assessed.

With respect to stress acceleration, six studies on negative parenting behaviors, maternal depressive symptoms, and parental substance were in line with this model (Chaplin et al., 2019; Heitzeg et al., 2008; Heitzeg et al., 2010; Hulvershorn et al., 2013; Kwon et al., 2021; Wang et al., 2020). In the study examining parenting behavior, a mediation model was not conducted, as there were no significant associations between brain function and externalizing behavior. While the studies examining parental depressive symptoms and parental substance use did not assess for mediation, two of the five studies demonstrated significant associations between frontolimbic activation/connectivity and externalizing behavior (Heitzeg et al., 2010; Wang et al., 2020).

Interestingly, some results were not accounted for within the models examined in this review. For example, disruption in frontoparietal regions, rather than in frontolimbic regions, was noted for studies examining threat-based parenting behaviors (i.e., corporal punishment, physical maltreatment; Barbosa et al., 2018, Shackman & Pollak, 2014). However, the proposed mediation model was not supported in these studies.

It was noted that neurodevelopmental patterns differed across studies examining the same or similar parental constructs. For example, in some cases biological sex may moderate these associations, as threat-related parenting behaviors were associated with altered frontoparietal activation in females, while threat-related parenting supported accelerated frontolimbic development among males. More longitudinal work is needed to assess whether this accelerated frontolimbic development in males is adaptive for youth, as early development in a given region may result in later structural underdevelopment (e.g., smaller volume; Deoni et al., 2016; Shaw et al., 2006). Similarly, there was sex-based variation observed in early childhood resting-state functional connectivity among youth whose mothers endorsed postnatal maternal depressive symptoms. Females exhibited altered frontolimbic functioning (i.e., difficulties with emotion processing and regulation), more consistent with the dimensional threat model, whereas males exhibited altered frontoparietal functioning (i.e., difficulties with attention to emotional stimuli), in line with the dimensional deprivation model. Thus, more studies examining biological sex as a moderator are needed to better understand how brain structure and function is altered in each sex.

Regarding findings on frontal brain volume, Demers and colleagues (2019) did not find strong evidence of accelerated brain development due to maltreatment history (i.e., threat) as expected based on prior work on the stress acceleration model (Callaghan & Tottenham, 2016). However, there was a marginal association identified between maltreatment history and higher frontal lobe volume for those who experienced only one type of maltreatment, as opposed to multiple forms of maltreatment. Thus, accelerated development of frontal areas may be dependent upon the number of and types of maltreatment exposure. It is important to note that incidents of both abuse and neglect, inclusive of but not restricted to a parent, were captured under maltreatment in this study; therefore, it is not clear whether this marginal effect was related to deprivation- or threat-related parental factors. Further, neuroimaging for this study was conducted in adulthood, at which point there may be developmentally appropriate reductions in frontal volume (Peters, 2006), limiting our knowledge on whether significant effects would have been observed earlier in adolescence.

In some studies, youth who had parents with substance use disorders demonstrated evidence of accelerated or typical frontolimbic development via higher frontal activation and lower limbic activation, consistent with the stress acceleration model. Prior work has shown that having a parent who engages in substance misuse can be considered a traumatic event (Parolin et al., 2016), potentially providing a similar maltreatment (i.e., threat) context as those youth demonstrating accelerated development of frontolimbic neurocircuitry. In contrast, two studies that included youth from middle childhood to young adulthood found smaller volumes in frontal regions and another study found lower frontal brain activation (i.e., delays in neurodevelopment), consistent with the dimensional model. In the latter study, the youth were preadolescents and thus may not yet have gone through normative PFC development. Given the wide age range, it is difficult to discern whether these findings are related to normative development prior to adolescence or suggest heterogeneity in structural brain outcomes of youth with parents who have history of substance use. Further, one study examined prenatal alcohol exposure, which may have different implications for brain development than a history of substance use. Notably, it is not possible to discern whether parents who used substances were more withdrawn or hostile in the reviewed studies. Kepple (2018) found that past year parental substance use disorder (SUD) diagnosis was associated with neglect, while past year substance use (i.e., not necessarily meeting criteria for SUD) was associated with abuse, suggesting differential pathways between substance use and parental deprivation or threat. Thus, heterogeneity in neural functioning may provide support for divergent pathways of neurodevelopment for youth of parents who have substance use difficulties. It is imperative for future work to further break down the assessment of parental psychopathology, examine parent-child associations longitudinally, and take pubertal factors into account in order to parse normative and non-normative development as well as resultant behavioral outcomes.

In sum, most studies demonstrated support for the dimensional model, while a more limited number provided support for the stress acceleration model, when considering both parental threat- and deprivation-contexts. Although parental behaviors and psychopathology consistently predicted youth externalizing behaviors in the expected directions, there was only limited overall support for the proposed mediational model, given that many of the reviewed studies did not explicitly test for mediation. Further, several findings fell outside of the proposed model (i.e., frontoparietal disruption in a threat context) and may suggest heterogeneity within parental factors.

## Limitations

These findings may be limited as the existing literature primarily relied on community samples, which may have reduced the ability to identify significant brain-behavior associations due to low levels of negative parenting behaviors and youth symptoms. As such, examination of clinical samples will be important in future studies to identify the most at-risk youth. Additionally, conclusions were limited in the thorough examination of brain-behavior associations across parents and youth, as well as the examination of biological sex as a potential moderator, as some reviewed studies solely included either parents or children of one biological sex. Additionally, it was difficult to draw concrete conclusions about whether neural functioning was normative or delayed for a given sample, as not all reviewed studies considered hormonal development. Further current study limitations involved the reliance of most reviewed studies on a single assessment method for parental factors (e.g., observational vs. self-report, categorical diagnoses vs. continuous symptoms) and much variation across the task methodologies across studies (e.g., use of facial expressions versus emotional valence pictures) and neuroimaging (e.g., EEG, rs-fMRI, task-based fMRI). For example, reviewed studies may have produced divergent findings as a result of varied task demands or reporter biases rather than true differences related to parental deprivation or threat. Lastly, it was not possible to thoroughly assess changes in brain development and behavior within this study, as many of the reviewed studies were cross-sectional or were underpowered to detect a significant mediation effect due to small sample size.

### Call to Action for More Research on Parental Factors and Youth Neurocircuitry

This review sheds light on the differential associations between parental factors characterized by deprivation and threat and on youth frontolimbic and frontoparietal neurocircuitry, as well as externalizing behavioral functioning. However, conclusions should be interpreted with caution given the limited number of studies, as well as the limitations in the existing literature. There is a need for more research to fully understand the development of neurocircuitry involved in emotion regulation, as well as the etiology of externalizing behaviors. The following recommendations for future research may lead to a more comprehensive understanding of this development, which is crucial to improvements in the provision of effective and efficient clinical care to families.

1. **Research with community-based and clinical samples**. With the exception of substance use disorders, most of the studies included in this review solely examined effects within community samples. While this approach may be more reflective of the general population, low baseline levels of parental and/or youth symptoms may make it difficult to identify significant parent-child effects. Future studies should recruit clinical samples and investigate neural and behavioral differences in comparison to healthy control groups. Additionally, parental and youth psychopathology should be examined both categorically and dimensionally in order to capture effects on youth functioning within families of those parents and youth presenting with subthreshold presentations.
2. **More clarity in reporting and examining parental symptoms and/or behaviors**. Within the current review, findings on parental depression and parent substance use aligned with both deprivation- and threat-based pathways. These differences may suggest heterogeneity in symptomatology and/or clinical presentation within these disorders. Thus, future studies should provide more information on the specific presenting symptoms within samples and conduct analyses using groups of deprivation-versus threat-based symptoms to better examine these constructs. Further, terms such as “maltreatment” are more general and may refer to neglect (i.e., deprivation) or physical abuse (i.e., threat). Thus, these terms should be well-defined within studies and separated in order to accurately test pathways involving parental factors characterized by deprivation and threat.
3. **Examining moderators of effects (age, biological sex, pubertal development)**. In addition to parental factors, several studies investigated relevant child factors such as age, biological sex, and pubertal development, while identifying significant effects on neural functioning. In some studies, these factors were limited by inclusion/exclusion criteria, and thus, it was not possible to determine whether there were moderation effects due to these factors. Given their importance in shaping youth neurodevelopment, these factors should consistently be examined in studies of youth neural structure and function. More specifically, future work should aim to look at changes in brain-behavior associations across time, while incorporating these developmentally-relevant factors.
4. **Examining constructs using multi-method assessment**. The studies in this review tended to include only one method (e.g., self-report, observational) of assessing parental factors. Future studies should consider the use of multi-method assessment for parental factors in order to minimize potential reporter biases and examine consistencies in findings, or lack thereof, across assessment methods. For example, parents with psychopathology may be less accurate reporters of their own symptoms, and thus an observational measure may be more valid. Further, multi-method assessment should be used to validate constructs (e.g., emotion regulation) via both subjective (e.g., self-report) and objective (e.g., neuroimaging) measures.
5. **Examining effects of parenting on executive attention vs. emotion regulation neural networks**. The dimensional model, as well as the newly presented model in this review, showed distinctions between the neural networks that manage executive attention to emotional stimuli (i.e., frontoparietal) and emotion regulation (i.e., frontolimbic). For example, the threat-based pathway showed alterations in the frontolimbic emotion regulation network, while the deprivation-based pathway showed alterations in the frontoparietal executive attention network. Yet some reviewed literature showed evidence of altered frontoparietal functioning (i.e., lower frontoparietal activation) with exposure to threat-based parental factors, and altered frontolimbic functioning (i.e., higher limbic and lower frontal activation) with exposure to deprivation-related parental factors. By purposefully investigating these pathways separately through use of network parcellations, nuanced differences in how parental factors impact each pathway and how altered functioning may affect youth behavior can be observed.

## Concluding Remarks

Overall, this review demonstrated that parental factors characterized by deprivation and threat are differentially associated with frontolimbic and frontoparietal structure and function in youth with and without externalizing behaviors. There was strong evidence to support the dimensional model, as well as some evidence to support the stress acceleration model with respect to the deprivation and threat context. Several studies examining parental depressive symptoms supported the newly proposed mediation model; the other reviewed studies either did not support this model or did not assess for mediation. More work is needed to fully conceptualize deprivation-versus threat-based factors among parents who experience depressive symptoms and have a history of substance use. Building from extant models of neurodevelopment, there was evidence of disruption in both frontolimbic and frontoparietal networks across deprivation- and threat-based factors, rather than disruption within just one network. More specifically, findings of this review extended beyond existing models to include frontoparietal alterations with respect to threat and frontolimbic alterations with respect to deprivation. Further, variations by biological sex and pubertal factors played a role in neural functioning. This review partially supported prior notions that youth exposed to threat-related and deprivation-related parental factors may demonstrate earlier frontolimbic maturation. In contrast to the stress acceleration model, some youth exposed to deprivation-related factors showed delayed maturation beyond developmentally normative periods. By calling attention to these differences, we can better predict risk and resilience trajectories of emotional development in youth. Additionally, this knowledge may better inform individualized targets (e.g., parenting, youth emotion regulation skills) in universal prevention and clinical intervention efforts. It will be especially important to involve parents who endorse their own maladaptive parenting behaviors and/or psychopathology in the treatment of youth. Further, studies should follow up on whether treatments that specifically target youth emotion regulation difficulties lead to changes in brain development (e.g., maturation of frontolimbic and frontoparietal pathways) and reductions in behavioral difficulties in youth. For example, dialectical behavior therapy, which specifically targets emotion dysregulation for both parents and children, has shown promise among adolescents and adults with externalizing disorders (Abootorabi Kashani et al., 2020; Bayat et al., 2020; Meyer et al., 2020). Overall, the current review provides a synthesis of the extant literature on parental factors and youth neural functioning and behavior, as well as a model from which to examine forthcoming results.

## Data Availability

All data reviewed is contained in the manuscript and further details may be found in the respective articles cited within this review.

## Acknowledgments

Support for this project was provided by the National Institute of Mental Health (K23 MH117280-01A1).

## Author Contributions

KEF conceived and designed the project. KEF, RPL and MMH conducted literature search procedures. KEF wrote the paper and all authors contributed to the revisions and approved the final version.

